# Neural network-based identification of easily-obtainable demographic and clinical characteristics to identify people with tuberculosis

**DOI:** 10.1101/2025.10.23.25338536

**Authors:** Devendra Singh Parihar, Joshua Jansen van Vüren, Thomas Niesler, Grant Theron, Daphne Naidoo, Marisa Klopper, Frank Cobelens, Lutz Kolbe, Kimsey Zajac, Willy Ssengooba, Moses Joloba

## Abstract

We consider the application of machine learning to the classification of tuberculosis (TB) based on clinical and demographic data. Such data is routinely collected from people who present with cough at community-level care centres. Therefore, such automatic classification could identify people who require expensive but critical confirmatory testing, thereby offering simple and low-cost method of triage. Logistic regression, XGBoost, and convolutional neural network classifiers are evaluated using fully-nested cross validation, with and without feature selection. Although the application of CNNs to clinical and demographic data is unconventional, we show it to be effective. Experiments are carried out using two datasets: cough diagnostic algorithm for TB (CODA TB), *n* = 1140 and cough audio triage for TB (CAGE-TB), *n* = 463, for both datasets all participants self-presented to healthcare facilities with symptoms or risk factors suggestive of TB. Using the CNN, areas under the receiver operating characteristic (AUROC) of 80.48% and 83.06% are achieved for the two datasets respectively. Furthermore, performance is shown to improve both when the set of clinical features is extended, and when the number of people in the dataset increases. This holds promise of the development of an automated TB triage tool, implemented on a low-cost mobile device such as a smartphone, that is suitable for use at primary health-care facilities.

## 1 Introduction

In 2023, tuberculosis (TB) resulted in an estimated 1.25 million deaths worldwide [World Health Organization, 2024]. An important contributing factor to this persistently high figure is the large number of people that remain undiagnosed and therefore continue to transmit TB. This is a particular concern in low and middle-income countries, where access to confirmatory diagnostic tests for TB is limited, especially in community-based or decentralised health care centres [Huddart et al., 2016, Nathavitharana et al., 2019]. Triage-based approaches can address this by offering an accessible, scalable and low-cost approach to decrease the number of people who must undergo subsequent expensive confirmatory testing, while also minimising the number of TB cases that are not detected.

Bartl et al. [2025] have developed and validated a random forest model to predict whether an individual with HIV develops TB within 6 months from the moment the biomarkers were collected. This model was trained on 55 people with HIV (PWH) developed TB and 1432 PWH who did not get TB, using 48 demographic, clinical, and lifestyle variables. The model achieved predictive performance, with an AUROC of 83%, a sensitivity of 70.1%, and a specificity of 81.0%. Even this baseline model, using the top 20 features, maintained acceptable performance (AUROC 74%) and showed generalizability across datasets. While the model showed strong performance, its predictive accuracy relies on the biomarkers from PWH.

Orjuela-Cañón et al. [2025] investigated the use of natural language processing and machine learning techniques to support pulmonary tuberculosis (PTB) diagnosis in low-resource settings. The study used data from 233 patients treated at the Hospital Santa Clara, in Colombia, comprising structured clinical data and unstructured physician reports. Five machine learning models were evaluated: support vector machines, *k*-nearest-neighbour classifiers, logistic regression, random forests and artificial neural networks. Some of these used the clinical data, others the physician reports, and some a combination of both through data fusion strategies. The best clinical data-based model was a neural network, achieving an AUROC of 69.9%. The physician reports-based models, despite lower specificity, showed higher sensitivity (73%). Data fusion approaches, particularly classifier stacking, improved sensitivity (71.3%) but showed only moderate overall performance. This study utilizes a relatively small dataset, with clinical data restricted mostly to demographic characteristics.

Chen et al. [2024] have analysed longitudinal electronic health record data gathered from 4,540 patients between 2017 and 2021 at the National Clinical Research Center for Infectious Diseases in Shenzhen, China, to identify TB in HIV-positive individuals. A multi-layer perceptron neural network, using 48 clinical variables as input, achieved modest performance (AUROC: 61.6-68.2%). In contrast, a long short-term memory (LSTM) recurrent neural network using only laboratory test data as input demonstrated stronger predictive capability (AUROC: 82.3-85.0%), outperforming electronic health record only models. The LSTM also showed consistent results in 5-fold cross-validation, indicating its robustness for detecting HIV-TB co-infections. Since the study focused on a population of HIV-positive patients, generalisability and clinical utility across diverse populations is a limitation.

Luo et al. [2022] aimed to distinguish between active tuberculosis and latent tuberculosis infection using standard clinical laboratory tests, including complete blood count (CBC), biochemical, coagulation, and inflammatory indicators. Data from 892 participants (discovery cohort) and 263 participants (validation cohort) were used to develop 28 machine learning models. Among these, 25 achieved a test set AUROC above 90%. Best performance was achieved using conditional random forests, with an AUROC of 97.8%, a sensitivity of 93.4%, and a specificity of 91.2%. Although this study achieved high performance in distinguishing active tuberculosis from latent infection, their study relied only on routine laboratory test indicators. This dependence on standard laboratory tests restricts applicability in resource-limited settings.

Ghermi et al. [2024] introduce TubIAgnosis, a machine learning web application designed to assist the diagnosis of active tuberculosis (TB) using demographic and clinical data, specifically CBC parameters, age, and sex. Eighteen CBC-derived features, along with patient age and sex, were used to train various classifiers. Algorithms such as logistic regression, random forest, gradient boosting, XGBoost and others were evaluated. Among these, the SMOTE-enhanced XGBoost model delivered the best performance, achieving an AUROC of 90.6%, surpassing the balanced XGBoost model (AUROC = 89.4%). The model’s performance may be affected by the heterogeneity in TB forms (pulmonary vs extrapulmonary) and varying patient severity profiles. Moreover, the model’s performance on unbalanced data remains to be investigated.

Botha et al. [2018] used logistic regression to classify TB in a dataset consisting of 17 TB-positive patients and 21 healthy controls. Five clinical measures: mid upper arm circumference, temperature, body mass index (BMI), anaemic conjunctivae and heart rate, were used to train a classifier, achieving an AUROC of 80%, a sensitivity of 72% and a specificity of 69%. It should be borne in mind, however, that this work was based on a small dataset, and distinguished between TB positive patients and healthy controls.

Thus, work considering the application of machine learning to the detection of TB based on demographic and clinical data has so far either focused on specific cohorts (such as HIV-positive patients) or has relied on standard laboratory test indicators. Furthermore, in many cases the datasets employed have been limited in size or were collected in a hospital setting. These factors limit the generalisability of the results.

In this study, we consider TB detection by machine learning based on a set of easily-accessible demographic and clinical data. Furthermore, we base experiments on two diverse and fairly large datasets that include symptomatic people from several countries and that were collected at community health centres. We conduct experiments utilising these two datasets both in isolation and also in combination. Finally, we apply a convolutional neural network (CNN), an architecture that is not conventionally associated with this type of data, to the classification problem and demonstrate improved performance. This performance is not far from the 60% specificity at 90% sensitivity which recently was the target product profile recommended by the WHO for community-based triage using a two-step screening test World Health Organization [2025].

## 2 Data

We have employed two datasets for this work, brief descriptions of which are presented below. Both of these datasets comprise people who have self-presented to a community health care centre with symptoms which are suggestive for TB. Therefore, the composition of these cohorts may differ from a random sample of the population.

### 2.1 The CODA TB dataset

This publicly available TB diagnostic dataset was compiled for use in CODA TB Dream Challenge [SAGE Bionetworks., 2024, Jaganath et al., 2025]. It includes clinical data collected from 1140 people across seven countries: Uganda, the Philippines, Vietnam, Madagascar, India, South Africa, and Tanzania [Huddart et al., 2024]. For each person, a set of 15 clinical and demographic features is provided, as listed in Table 1.

**Table 1:** Demographic and clinical data gathered for each person in the CODA TB and CAGE-TB datasets. For CODA all variables, except height, weight, heart rate and temperature, were self-reported.

| Feature | Available for |  | Availability class |
| --- | --- | --- | --- |
|  | CAGE-TB | CODA TB |  |
| Location of clinic | ✓ |  | A |
| Arthritis | ✓ |  | A |
| Age | ✓ | ✓ | A |
| Dyspnea | ✓ |  | A |
| Sex | ✓ | ✓ | A |
| Chest pain | ✓ |  | A |
| Prior TB | ✓ | ✓ | A |
| Prior TB type pulmonary |  | ✓ | A |
| Prior TB type extrapulmonary |  | ✓ | A |
| Night sweats | ✓ | ✓ | A |
| Fagerstrom score | ✓ |  | B |
| Fever within last two weeks | ✓ | ✓ | A |
| TBscoreII value | ✓ |  | B |
| Coughing up sputum | ✓ |  | A |
| Height | ✓ | ✓ | B |
| Hemoptysis |  | ✓ | A |
| Recent weight loss (>1.5kg) | ✓ | ✓ | A |
| Weight | ✓ | ✓ | B |
| Anaemic conjunctivae | ✓ |  | B |
| BMI | ✓ |  | B |
| CRP concentration | ✓ |  | C |
| Middle upper arm circumference | ✓ |  | B |
| Haemoglobin concentration | ✓ |  | C |
| Cough duration | ✓ | ✓ | A |
| Number of symptoms | ✓ |  | B |
| Covid status | ✓ |  | C |
| HIV status | ✓ |  | B |
| Current tobacco smoker | ✓ | ✓ | A |
| Axillary/ear temperature | ✓ | ✓ | B |
| Heart rate | ✓ | ✓ | B |
| Day of year | ✓ |  | A |
| Respiratory rate | ✓ |  | B |
| Years since previous TB | ✓ |  | A |
| Other chronic conditions | ✓ |  | A |
| Diabetic | ✓ |  | A |
| Hypertension | ✓ |  | A |
| Cancer | ✓ |  | A |
| Cholesterol | ✓ |  | A |
| Hyper/Hypothyroidism | ✓ |  | A |
| Asthma | ✓ |  | A |
| Antiretroviral therapy (ART) | ✓ |  | A |

Although BMI can be directly calculated from height and weight, it was elected to not include this value as input to the classifier in order to maintain some comparability to previous studies which employed the CODA TB dataset [Kafentzis et al., 2023]. Additionally, even though some included variables are derived from others (for example BMI from height and weight) we will show that including both often leads to an improvement in classification performance.

Of the 1140 people in the dataset, 293 were established to be TB positive and the remaining 847 TB negative using a microbiological reference standard in which a person is classified as being TB positive when Xpert MTB/RIF Ultra (Ultra; Cepheid, Sunnyvale, USA) tests positive for sputum or urine, when liquid or solid culture tests positive, when Ultra is positive on contaminated liquid culture, or when there are two trace positive Ultra results. If no positive results were obtained for Xpert or Ultra and two negative liquid or solid cultures or one liquid and one solid negative culture were obtained, then the person was classified as TB negative. People who had only one Ultra trace result and no other positive Xpert or Ultra results were classified as indeterminate. This standard is presented in Supplementary Table 7a.

The 1140 people were partitioned into ten folds, ensuring an approximate balance between the number of positive and negative TB cases. This resulted in at least 28 TB positives and 84 TB negatives per fold. These folds were used for cross-validation, as described in Section 3.2.

### 2.2 The CAGE-TB dataset

The CAGE-TB dataset was compiled from people who self-presented to one of three community health care centres near Cape Town, South Africa, with an at least two-week long cough. For each person, a set of 38 demographic and clinical features was collected, as listed in Table 1.

Of the 463 people in the dataset, 86 were established to be TB positive and the remaining 377 TB negative using a composite reference standard of either a Ultra positive or a MGIT 960 liquid culture. In cases where a person was HIV-positive, two cultures were performed. Specifically, a person is considered to be TB positive if they are culture positive for Mycobacterium tuberculosis complex (MTBC) or if they have a culture-negative result but a positive Ultra result (see also Supplementary Table 7b). If the culture is negative and there is no positive Ultra result, they are classified as TB negative. In cases of contaminated sputum culture results, Ultra performed on the contaminated culture growth is used, and a positive result is used as a TB positive classification. For any non actionable (not positive or negative) culture results, no cultures performed, or contaminated cultures without a positive Ultra, the result is considered unclassifiable.

The 463 people were partitioned into ten folds, ensuring an approximate balance between the number of positive and negative TB cases. This resulted in at least 8 TB positives and 37 TB negatives per fold. These folds were used for cross-validation, as described in Section 3.2.

### 2.3 Availability class

Each clinical or demographic feature was assigned to one of three availability classes, based on the level of ease with which the corresponding data can be obtained. Class A features are those that are self-reported as part of a single question or are readily available without requiring any counting or scoring. Class B features are those that require measurement and some degree of training or calculation by the health care professional. Finally, Class C features are those that require a test, which could be point of care or laboratory. The class which each feature belongs to is indicated in Table 1.

## 3 Methods

The following describes how our data was prepared before being presented as input to a classifier for training and evaluation. Each dataset is prepared separately but in the same way. This allows separate but comparable experiments per dataset, and also combined experiments using a pooled dataset.

### 3.1 Missing values

Instead of removing people with incomplete data, or attempting to artificially generate the values for example by interpolation, we elected to include an additional associated binary input for each such item that denotes whether the corresponding value is missing or available. Using this scheme, people with incomplete information could remain in the dataset. For example, if a person’s haemoglobin concentration was not measured, instead of excluding that person from our analysis, we set the corresponding unavailability indicator to TRUE, and the value for haemoglobin to zero. In the CAGE-TB dataset, 63 people do not have a measured haemoglobin value, and 88 do not have a recorded c-reactive protein (CRP). A complete list of the number of available datapoints is presented in Supplementary Table 6.

### 3.2 Nested k-fold cross validation

For both datasets, nested ten-fold cross validation was used to allow robust parameter training, hyperparameter optimisation and independent testing. Parameters refer to the values that are directly optimised during training, such as the weights used by logistic regression. Hyperparameters refer to other variables, such as the degree of regularisation, that are not determined during training but must also be optimised. Further information is provided in Supplementary Section 7.2.4. Each of the ten folds described in Section 2 is held-out in turn as a final test set, while the remaining nine are used for training and development. This *outer loop* results in ten classifiers with associated test-set scores, allowing the mean and standard deviation to be computed. Of the nine folds not used for testing, each is held out in turn to serve as a development set, while the remaining eight are used for training. This *inner loop* is used to optimise the hyperparameters. All data from a particular person resides in one of the ten folds to ensure that there is always complete separation between training, development and test sets.

### 3.3 Sequential Feature Selection (SFS)

For the CODA TB and the CAGE-TB datasets, 15 and 38 clinical and demographic features are available respectively (Table 1). However, not all of these features are necessarily relevant to the task of TB classification. Features that are not relevant can degrade classification performance when training data is limited, as it is for the work presented here. Our experiments include SFS, a method that iteratively evaluates feature subsets to identify those most relevant to a particular predictive model. This method is particularly effective in scenarios where the feature space is high-dimensional and domain knowledge about feature importance is limited [Rückstieß et al., 2011]. A detailed presentation on the procedure employed during SFS can be found in Supplementary Section 7.1.

### 3.4 Considered classifier architectures

We evaluated three distinct classifier architectures, logistic regression, XGBoost and a one-dimensional convolutional neural network (CNN). Logistic regression served as a baseline, offering a simple and interpretable linear model that approximates the posterior probability of the positive class. Although limited in its ability to capture complex interactions, it provides a useful reference for comparing the performance of more advanced models. XGBoost is a tree-based ensemble method that iteratively builds a sequence of weak classifiers, each trained to reduce the residual error of the ensemble. This architecture is well-suited to tabular data and has demonstrated strong performance in clinical settings [Ghermi et al., 2024]. Finally, we considered a CNN configured for one-dimensional input. Although CNNs are traditionally used for spatially structured data such as images, we adapted this architecture to explore its utility for structured clinical features.

Each of the considered classifiers has hyperparameters which are optimised through nested cross-validation. A more complete description of each architectures and its hyperparameters is provided in Supplementary Section 7.2.

### 3.5 Merged CODA TB and CAGE-TB dataset

In addition to separate experiments for each of the two datasets, we leverage the CODA TB dataset as additional training data by pooling it with the CAGE training data. The resulting classifiers are evaluated only on the CAGE development and test sets presented in Section 2.2. Since the clinical and demographic data gathered for CODA TB and for CAGE-TB differ, the data pooling necessitates the use of a set of features common to both datasets. From Table 1 we can identify a total of 12 such common features: sex, age, height, weight, duration of cough, prior TB, fever, night sweats, heart rate, temperature, weight loss and recent smoking. Since the feature set is small and the application of SFS for CODA TB had mixed results, we do not apply feature selection in these experiments, but employ the full set of 12 features.

## 4 Results

### 4.1 CODA TB dataset

An analysis of the SFS feature selection process for the CODA TB data is presented in Figure 1. The graph shows the average improvement afforded by each successively identified feature relative to chance (AUROC = 50%).

**Figure 1:**
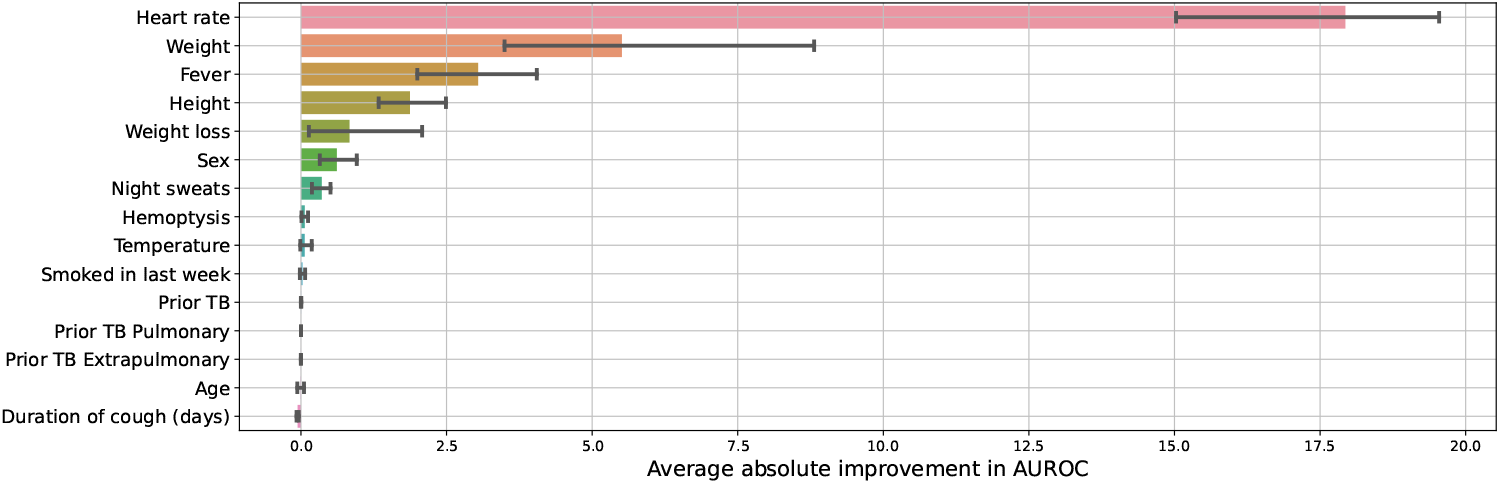
Per-feature average absolute improvement in development set AUROC during SFS for CODA TB, relative to chance (50% AUROC). Error bars indicate the bootstrap estimate of the 95% confidence interval across the development sets.

Heart rate is most informative, followed by weight, fever and height. Six features (smoking, prior TB, prior TB pulmonary, prior TB extrapulmonary, age and duration of cough) either provide negligible improvement or lead to a decrease in development set AUROC. An extended per-country analysis is provided in Supplementary Figure 5.

Table 2 presents the performance, in terms of AUROC, of the three considered classifiers, for both datasets, and both with and without SFS. For CODA TB, we see that, when no SFS is performed and the models are trained on the full set of 15 features, the CNN performs best on both development and test sets. However, when the 9 features identified by SFS are used instead, the best performance is achieved by logistic regression. We believe that one reason for this decrease is the required minimum size of the CNN kernel for a specific input vector, which may have negatively impacted model performance.

**Table 2:** Development and test set AUROCs for classifiers trained and tested on either the CODA TB or the CAGE-TB datasets, both when using all features (15 and 40 respectively) and when using the features identified by SFS.

| Dataset | Model | All features (no SFS) |  | Optimal features (with SFS) |  |
| --- | --- | --- | --- | --- | --- |
|  |  | Development | Test | Development | Test |
| CODA TB | LR | $80.37 \pm 0.61\%$ | $80.26 \pm 4.88\%$ | **$80.27 \pm 0.39\%$** | **$80.29 \pm 4.01\%$** |
| | XGBoost | $77.62 \pm 0.70\%$ | $78.02 \pm 3.52\%$ | $74.51 \pm 0.62\%$ | $74.99 \pm 4.07\%$ |
| | CNN | **$80.51 \pm 0.46\%$** | **$80.48 \pm 3.14\%$** | $79.72 \pm 0.53\%$ | $79.81 \pm 4.03\%$ |
| CAGE-TB | LR | $77.34 \pm 1.30\%$ | $77.01 \pm 9.51\%$ | $82.13 \pm 1.18\%$ | $81.56 \pm 7.49\%$ |
| | XGBoost | $80.43 \pm 1.79\%$ | **$82.48 \pm 7.24\%$** | $79.60 \pm 1.83\%$ | $79.39 \pm 7.37\%$ |
| | CNN | **$80.51 \pm 0.93\%$** | $80.40 \pm 8.76\%$ | **$83.14 \pm 1.05\%$** | **$83.06 \pm 7.17\%$** |

### 4.2 CAGE-TB datatset

An analysis of the SFS feature selection process for the CAGE-TB dataset is presented in Figure 2. The graph shows the average improvement afforded by each successively identified feature relative to chance (AUROC = 50%).

**Figure 2:**
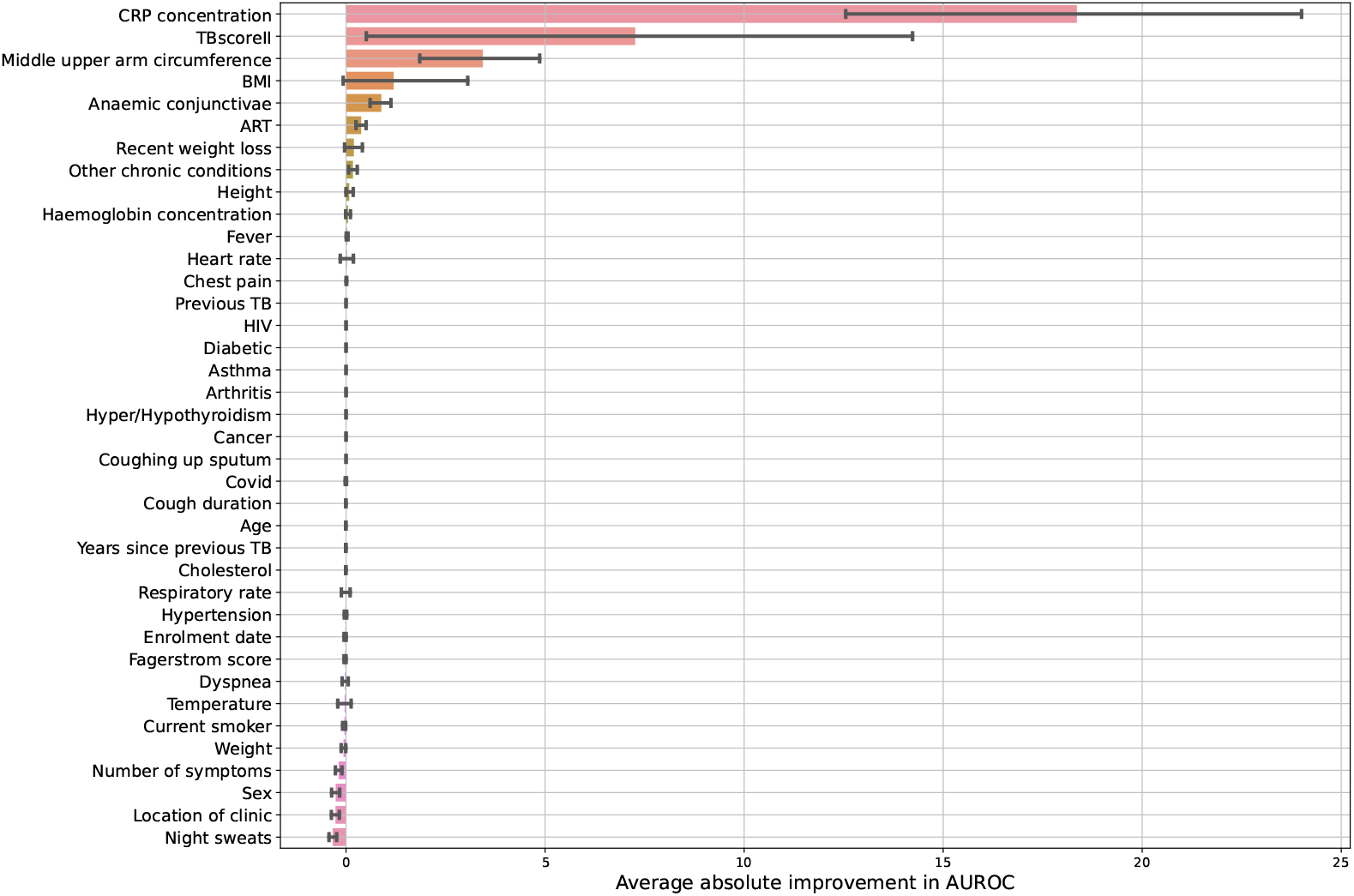
Per-feature average absolute improvement in development set AUROC during SFS for CAGE-TB, relative to chance (50% AUROC). Error bars indicate a bootstrap estimate of the 95% confidence interval across the development sets.

In contrast to the results for CODA TB (Figure 1) where heart rate was the overall most informative feature, for CAGE-TB it is ranked twelfth. Instead, CRP concentration, followed by TBscoreII, middle upper-arm circumference and BMI are the most informative features.

Furthermore, Table 2 shows that, when the models are trained on the full set of 38 features, the CNN performs best on the development set, but not on the test set, where XGBoost exhibits a higher AUROC. However, when the 13 features identified by SFS are used instead, the best performance on both development and test sets is achieved by the CNN. This is also the best performance overall for CAGE-TB.

Table 3 presents the development and test set AUROCs for the LR and CNN classifiers when input features are grouped by availability class (A only, B only, or A and B in combination). When using only the self-reported values (A), the LR and CNN classifiers achieve 67.16% and 71.62% test set AUROCs respectively. This improves to 73.18% and 75.88% respectively when using only those self-reported features identified during SFS. Utilising only values chosen by SFS from availability class B, which require some measurement to be taken or some basic intervention by a health care worker, further improves the test set AUROCs to 80.19% and 80.15% respectively. Finally, allowing SFS to choose from any feature in either availability class A or B (A+B) results in a further improvement, with the CNN achieving an AUROC of 82.18%. Further detail on the SFS results in these experiments is presented in Supplementary Section 7.4.

**Table 3:**
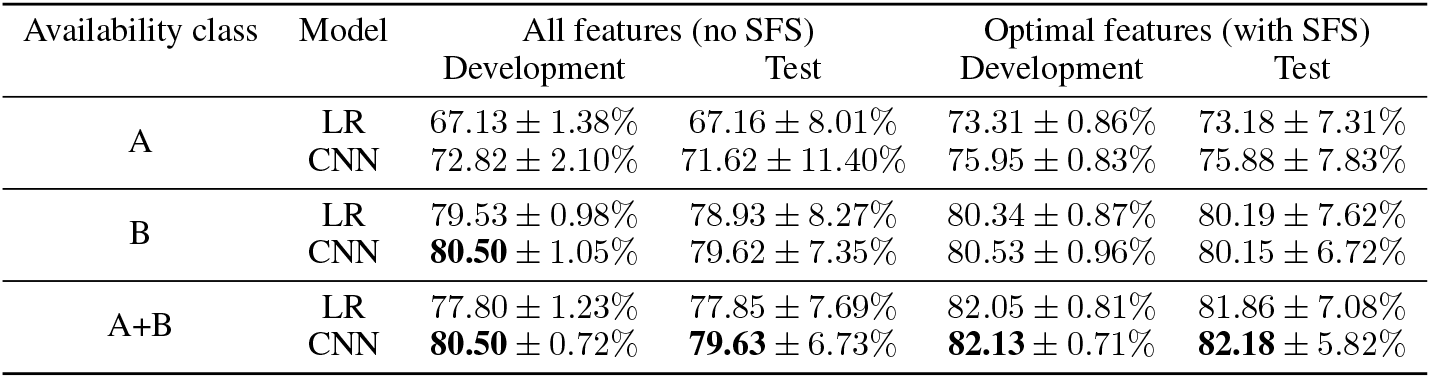
Development and test set AUROC for classifiers trained on the subsets of features separated according to the ease of availability (Table 1) class both on the entire subset and after further optimising the input features using SFS.

| Availability class | Model | All features (no SFS) |  | Optimal features (with SFS) |  |
| --- | --- | --- | --- | --- | --- |
|  |  | Development | Test | Development | Test |
| A | LR | 67.13 ± 1.38% | 67.16 ± 8.01% | 73.31 ± 0.86% | 73.18 ± 7.31% |
| | CNN | 72.82 ± 2.10% | 71.62 ± 11.40% | 75.95 ± 0.83% | 75.88 ± 7.83% |
| B | LR | 79.53 ± 0.98% | 78.93 ± 8.27% | 80.34 ± 0.87% | 80.19 ± 7.62% |
| | CNN | **80.50** ± 1.05% | 79.62 ± 7.35% | 80.53 ± 0.96% | 80.15 ± 6.72% |
| A+B | LR | 77.80 ± 1.23% | 77.85 ± 7.69% | 82.05 ± 0.81% | 81.86 ± 7.08% |
| | CNN | **80.50** ± 0.72% | **79.63** ± 6.73% | **82.13** ± 0.71% | **82.18** ± 5.82% |

### 4.3 Merged CODA TB and CAGE-TB

Table 4 presents the classification performance achieved when merging the CODA TB data with the CAGE-TB training sets, while optimising and testing on the CAGE-TB development and test sets. Classifiers trained only on the CAGE-TB data are included for comparison. Since these experiments use the reduced set of 12 features that are common to both datasets, the classifiers trained on only the CAGE data show reduced performance relative to Table 2, as may be expected.

**Table 4:** Development and test set AUROCs for classifiers trained on the merged CODA TB and CAGE-TB datasets, and tested on the CAGE-TB test set. All experiments use the full set of 12 features common to both datasets presented in Section 4.1. The performance of classifiers trained only on CAGE-TB is provided for comparison.

| Classifier | Trained on | Development | Test |
| --- | --- | --- | --- |
| LR | CAGE-TB | 73.87 ± 1.17% | 73.43 ± 8.90% |
| LR | CODA TB and CAGE-TB | 76.91 ± 1.14% | 75.95 ± 10.30% |
| XGBoost | CAGE-TB | 74.01 ± 1.86% | 73.66 ± 9.06% |
| XGBoost | CODA TB and CAGE-TB | 72.73 ± 1.60% | 72.11 ± 10.37% |
| CNN | CAGE-TB | 73.12 ± 1.57% | 72.52 ± 9.54% |
| CNN | CODA TB and CAGE-TB | **78.65** ± **1.16**% | **77.28** ± **10.64**% |

**Table 5:** Demographic and clinical data description from both CODA TB and CAGE-TB datasets.

| Feature | Description | Data type |
| --- | --- | --- |
| Location of clinic | Clinic at which the person was recruited | Multinomial (1,2,3) |
| Arthritis | Indicates that the person suffers with a chronic condition caused by inflammation of one or more joints, causing pain and stiffness that can worsen with age | Binary (Y/N) |
| Age | The age of the person | Quantitative (years) |
| Dyspnea | Indicates that the person has been experiencing a shortness of breath | Binary (Y/N) |
| Sex | The biological gender of the person | Binary (M/F) |
| Chest pain | Chest pain | Binary (Y/N) |
| Prior TB | Self reported status of whether the participant had or been told to have TB | Binary (Y/N) |
| Prior TB type pulmonary | Self reported status of whether the participant had or been told to have pulmonary TB | Binary (Y/N) |
| Prior TB type extrapulmonary | Self reported status of whether the participant had or been told to have extrapulmonary TB | Binary (Y/N) |
| Night sweats | Night sweats | Binary (Y/N) |
| Fagerstrom score | Indicates the persons's level of nicotine dependence | Quantitative (-6,0,1,...,10) |
| Fever within last two weeks | If the person has experienced any fever in the past two weeks | Binary (Y/N) |
| TBscoreII value | Person should have one or more symptoms (prolonged cough, dyspnea, chest pain, anaemia, middle upper arm circumference, BMI) | Quantitative (1,2,...,8) |
| Coughing up sputum | Coughing up sputum (spit from deep in the lungs) | Binary (Y/N) |
| Height | Person's height in centimetres | Quantitative (cm) |
| Hemoptysis | Self reported status of whether the participant has ever coughed blood | Binary (Y/N) |
| Recent weight loss (>1.5kg) | Weight loss | Binary (Y/N) |
| Weight | Person's weight in kilograms | Quantitative (kg) |
| Anaemic conjunctivae | Indicates that the inner part of the person's lower eyelid looks pale, which can be a sign of low red blood cells or anaemia | Binary (Y/N) |
| BMI | Person's body mass index | Quantitative (kg/m<sup>2</sup>) |
| CRP concentration | A protein made by the liver and released into the blood in response to inflammation in the body | Quantitative (mg/L) |
| Middle upper arm circumference | Persons's arm circumference | Quantitative (cm) |
| Haemoglobin concentration | It is used clinically to determine the presence of anemia | Quantitative (g/dl) |
| Cough duration | The number of days the person has been experiencing a cough | Quantitative (days) |
| Number of symptoms | Number of symptoms experienced by the person (i.e., fever, night sweats, excessive weight loss) | Multinomial (1,2,3,4) |
| Covid status | Sars-CoV-2 test result | Binary (Y/N) |
| HIV status | Indicates if the person is living with HIV | Binary (Y/N) |
| Current tobacco smoker | Indicates if the person currently smokes tobacco products | Binary (Y/N) |
| Axillary/ear temperature | Measurement of the person's body temperature | Quantitative (Celsius) |
| Heart rate | Measure of heart beats of the person per minutes | Quantitative (BPM) |
| Day of year | Convert the person' enrolment date into the corresponding day of the year | Quantitative |
| Respiratory rate | Measurement of the number of breaths someone takes every minute | Quantitative (breaths/min) |
| Years since previous TB | Indicates the year that the person had TB previously | Quantitative (years) |
| Other chronic conditions | Indicates if the person has any conditions that last 1 year or longer | Binary (Y/N) |

| Feature | Description | Data type |
| --- | --- | --- |
| Diabetic | Indicates if the person has a condition caused by too high blood sugar levels | Binary (Y/N) |
| Hypertension | Indicates that the person suffers with high blood pressure | Binary (Y/N) |
| Cancer | Indicates that the person suffers from a disease caused by an uncontrolled division of abnormal cells in a part of the body | Binary (Y/N) |
| Cholesterol | Indicates that the person suffers with a high level of bad cholesterol | Binary (Y/N) |
| Hyper/Hypothyroidism | Indicates that the person's thyroid gland makes too much (hyperthyroidism) or too little (hypothyroidism) thyroid hormone | Binary (Y/N) |
| Asthma | Indicates that the person suffers with a chronic lung disease caused by inflammation and muscle tightening around the airways, which makes it harder to breathe | Binary (Y/N) |
| Antiretroviral therapy (ART) | Indicates if the person is on Antiretroviral therapy | Multinomial (No - HIV neg, No - HIV pos, Yes - HIV pos) |

**Table 6:** Availability of clinical and demographic data gathered for each person in the CODA TB and or CAGE-TB datasets. Variables with missing entries are denoted in bold.

| Feature | Availability CAGE | Availability CODA |
| --- | --- | --- |
| Location of clinic | 463 | - |
| Arthritis | 463 | - |
| Age | 463 | 1 140 |
| Dyspnea | 463 | - |
| Sex | 463 | 1 140 |
| Chest pain | 463 | - |
| Prior TB | 463 | 1 140 |
| Prior TB type pulmonary | - | 1 140 |
| Prior TB type extrapulmonary | - | 1 140 |
| Night sweats | 463 | 1 140 |
| Fagerstrom score | 463 | - |
| Fever within last two weeks | 463 | 1 140 |
| TBscoreII value | 463 | - |
| Coughing up sputum | 463 | - |
| Height | 463 | 1 140 |
| Hemoptysis | 463 | 1 140 |
| Recent weight loss (>1.5kg) | 463 | 1 140 |
| Weight | 463 | 1 140 |
| Anaemic conjunctivae | 463 | - |
| BMI | 463 | - |
| CRP concentration | **375** | - |
| Middle upper arm circumference | 463 | - |
| Haemoglobin concentration | **400** | - |
| Cough duration | 463 | 1 140 |
| Number of symptoms | 463 | - |
| Covid status | **460** | - |
| HIV status | 463 | - |
| Current tobacco smoker | 463 | 1 140 |
| Axillary/ear temperature | 463 | 1 140 |
| Heart rate | 463 | 1 140 |
| Day of year | 463 | - |
| Respiratory rate | 463 | - |
| Years since previous TB | **114** | - |
| Other chronic conditions | 463 | - |
| Diabetic | 463 | - |
| Hypertension | 463 | - |
| Cancer | 463 | - |
| Cholesterol | 463 | - |
| Hyper/Hypothyroidism | 463 | - |
| Asthma | 463 | - |
| ART status | 463 | - |

**Table 7:**
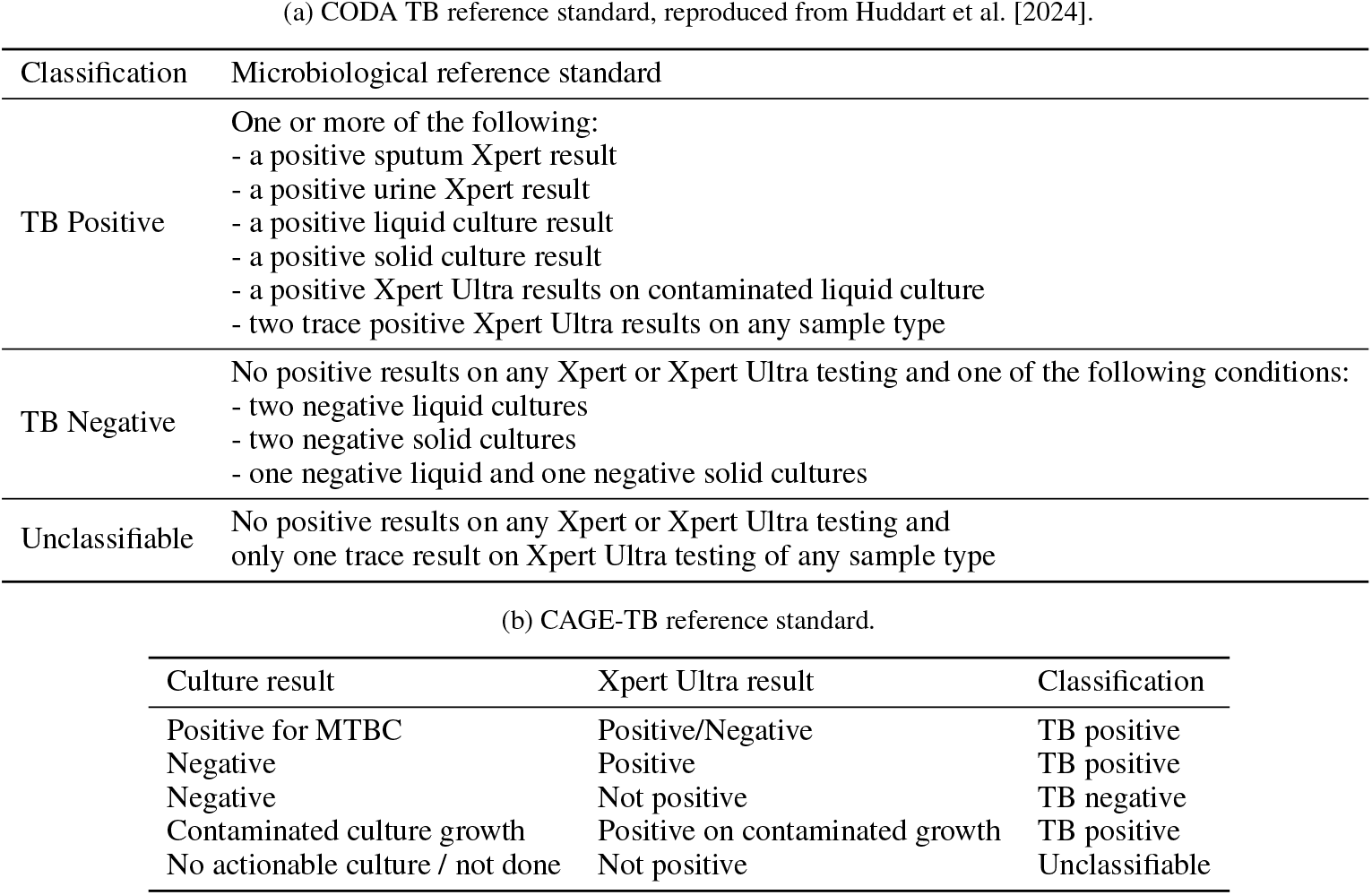
CAGE-TB and CODA TB reference standards.

(a) CODA TB reference standard, reproduced from Huddart et al. [2024].
| Classification | Microbiological reference standard |
| --- | --- |
| TB Positive | One or more of the following:<br>- a positive sputum Xpert result<br>- a positive urine Xpert result<br>- a positive liquid culture result<br>- a positive solid culture result<br>- a positive Xpert Ultra results on contaminated liquid culture<br>- two trace positive Xpert Ultra results on any sample type |
| TB Negative | No positive results on any Xpert or Xpert Ultra testing and one of the following conditions:<br>- two negative liquid cultures<br>- two negative solid cultures<br>- one negative liquid and one negative solid cultures |
| Unclassifiable | No positive results on any Xpert or Xpert Ultra testing and only one trace result on Xpert Ultra testing of any sample type |

(b) CAGE-TB reference standard.
| Culture result | Xpert Ultra result | Classification |
| --- | --- | --- |
| Positive for MTBC | Positive/Negative | TB positive |
| Negative | Positive | TB positive |
| Negative | Not positive | TB negative |
| Contaminated culture growth | Positive on contaminated growth | TB positive |
| No actionable culture / not done | Not positive | Unclassifiable |

Table 4 also shows that the additional CODA TB data improves the performance of all classifiers for both development and test sets, with the exception of XGBoost. For the CNN, the additional data improves the AUROC from 73.12% to 78.65% on the development set, and from 72.52% to 77.28% on the test set.

## 5 Discussion

When considering the results obtained separately for the two datasets (Table 2) we see that, when all features are used, the performance achieved for each dataset is fairly similar. For CODA TB, the CNN classifier achieves the best AUROCs on both the development and test sets. While the CNN also leads to the best development set AUC for CAGE-TB, this architecture does not deliver the best test set result. This may indicate overtraining, since CAGE-TB has almost three times as many features but fewer than half as many people as CODA TB.

When an optimal set of features is chosen by SFS, we see little change in the results for CODA TB. However, SFS results in a substantial improvement for CAGE-TB for both logistic regression and the CNN. In this case the CNN outperforms the other two architectures on both the development and test sets. We conclude that better generalisation is achieved for CAGE-TB by the smaller number of features chosen by SFS. We also see that, while the AUROCs were similar for CODA TB and CAGE-TB when the full features sets were used, the optimisation of the feature set by SFS affords the CAGE-TB systems a noticeable lead over the corresponding CODA TB systems. This indicates that the chosen subset of CAGE-TB features is more informative for TB classification than the similarly obtained CODA TB subset. This conclusion is supported by Figures 1 and 2, which reveal that the top six most informative features chosen for CAGE-TB are all not available for the CODA TB dataset.

We also see that, while the occurrence of night sweats was a useful feature in the CODA TB experiments, this feature was counterproductive in the case of CAGE-TB. While night sweats are a common symptom of TB, they are also a non-specific symptom that can be attributed to various other conditions, including hormonal imbalances, medication and infections other than TB. It could be the case that, in the CAGE cohort, such other factors were more pronounced than in the CODA cohort. Furthermore, the feature indicating whether a person is on ART is always selected before the feature indicating whether or not they are HIV positive. Knowing the presence of chronic conditions in isolation also does not seem to convey useful information with respect to TB classification. However, when grouped under the single indicator “other chronic conditions”, this value becomes useful. Surprisingly, for both datasets, prior TB, which is a known risk factor for TB, is not found to improve classification performance. We believe that this is because the prior TB indicator does not provide additional information when considered in addition to the variables already chosen by SFS (CRP, TBscoreII, BMI, anaemia, ART, recent weight loss, other chronic conditions, height, haemoglobin, fever, heart rate, and chest pain). These findings motivate further research into clinical measurements that can support a TB classifier.

We note that SFS is not effective for the XGBoost architecture for either CODA TB or CAGE-TB. This is however not surprising, as XGBoost is based on decision trees, which inherently perform feature selection by choosing those which provide best class separation. Therefore, additionally applying SFS is counterproductive.

It is noteworthy that, despite utilising exclusively the self-reported values in availability class A, the CNN classifier is able to achieve an AUROC which is only approximately 8% below that of our best performing classifier. Further, when SFS is applied to availability class A and B in combination, optimal performance is achieved when utilising a vector which includes only 12 inputs. Four of these are self-reported: ART status, recent weight loss, fever, and asthma, and the remaining eight some measurement or calculation (availability class B): TBScoreII, anaemic conjunctivae, height, middle upper arm circumference, BMI, heart rate, temperature, and HIV. The CNN classifier trained on this vector achieves an AUROC of 82.18% which is approximately 1% below the best performance achieved for CAGE-TB, despite never utilising inputs which are time-consuming or require special testing equipment to capture. Further discussion of the optimisation of the input vector for each of the availability classes is presented in Supplementary Section 7.4.

When the CODA TB data is pooled with CAGE-TB, the training sets are substantially enlarged. However, since only the 12 features common to both datasets could be used in these experiments, performance drops relative to systems using the optimal set chosen by SFS. Nevertheless, the results in Table 4 show that the additional data affords clear improvements in performance for logistic regression and the CNN. This indicates that a classifier based on clinical and demographic data can be improved by enlarging the dataset, and that this should therefore remain a key objective of ongoing work. Furthermore, while the AUROC achieved by the CNN using the 12 common features drops by approximately 5% absolute compared to the CNN trained only on the CAGE-TB data and using the 13 features identified by SFS, it nevertheless achieves an AUROC of 77.3% despite using only information that is easily accessible and does not require any POC testing. Further research that includes enlarging the dataset will tell whether additional performance improvements are possible using these same convenient 12 features.

The results in Tables 2 and 4 demonstrate the success of the CNN classifier. This success was surprising, since the clinical and demographic features lack inherent spatial relationships. We hypothesise that a favourable ordering of input features is able to place related information in close proximity and unrelated information further away. The fixed-dimension kernels in the first layer of the CNN can only capture patterns among these closely-located features, and completely disregards more distant relationships. By prohibiting the modelling of patterns between the distant and therefore unrelated features, noise in the form of spurious relationships is suppressed, which improves performance. The deeper layers of the CNN then combine these robust local relationships in order to make a final classification decision that takes all input features into account.

Recently, the WHO has recommended a minimum specificity of 60% at 90% sensitivity for triage testing World Health Organization [2025]. Figure 3 compares the ROCs of the LR, CNN and XGBoost classifiers for the subset of thirteen and nine clinical features selected by SFS for the CAGE-TB or the CODA TB datasets respectively. The performance for the full input vector is shown in Supplementary Figure 4.

**Figure 3:**
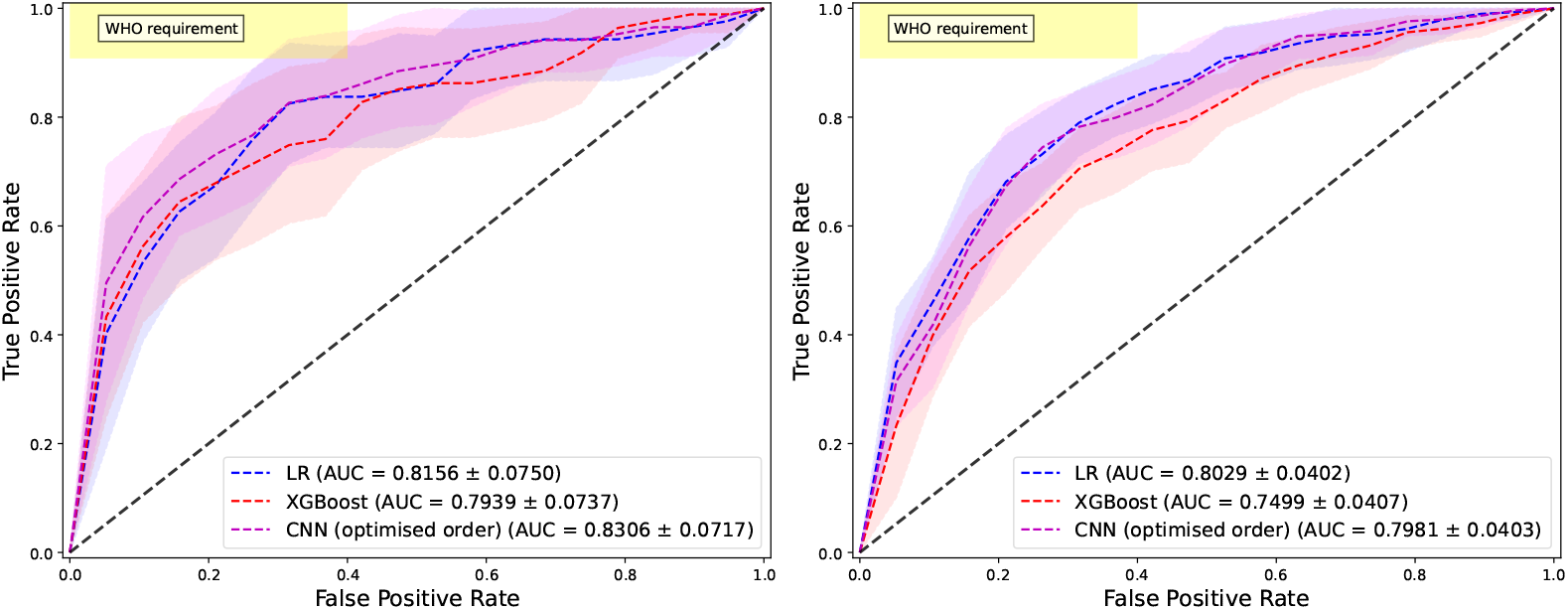
AUROC curves reflecting the performance of the logistic regression (LR), convolutional neural network (CNN), and XGBoost classifiers. Results are shown when using the subset of features identified by SFS for the CAGE-TB (left) and on the CODA TB (right) datasets.

**Figure 4:**
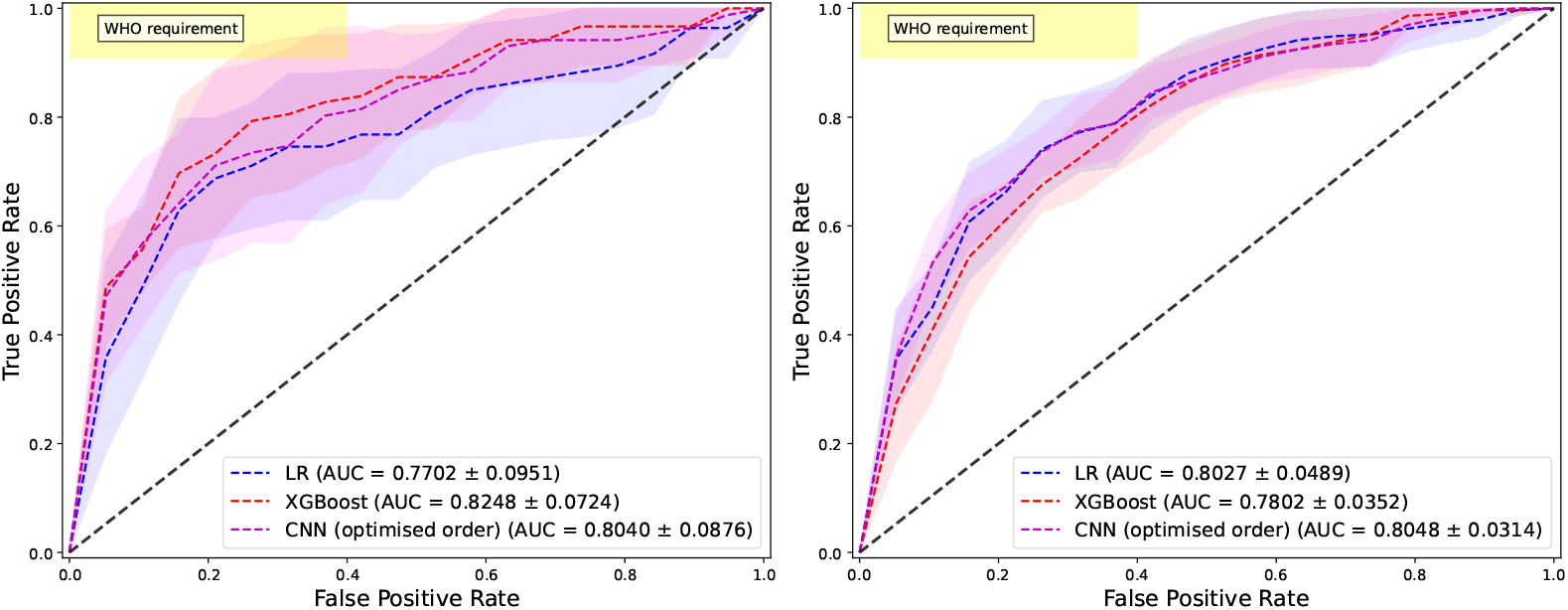
AUROC curves reflecting the performance of the LR, CNN, and XGBoost classifiers. Results are shown when using the full set of clinical data for the CAGE-TB (left) and on the CODA TB (right) datasets.

To achieve 90% sensitivity, the best specificity achieved by LR, XGBoost and the CNN is 42.11%, 26.32% and 47.37% respectively for CAGE-TB and 47.37%, 36.85% and 47.37% respectively for CODA TB. For the CAGE-TB dataset this falls short of the WHO benchmark of 60% by approximately 13%. However, our experimental results above indicate that gains can be achieved both by enlarging the pool of people and by collecting more clinical and demographic information. Therefore, data collection should remain high priority. Furthermore, it is perhaps possible that, with a dataset of sufficient size, the WHO targets can be fulfilled by a classifier requiring only easily-obtainable demographic and clinical input.

Finally, we would like to mention that the CODA TB dataset has been used previously for TB classification to develop a model incorporating both the clinical and demographic data we use here and recordings of cough audio Kafentzis et al. [2023]. The reported AUROC of this combined classifier was 81 ± 5%, but classification based only on the clinical and demographic features was not reported. Our results indicate that this classification performance, which uses both audio and clinical information, can in fact be achieved using the clinical data alone.

## 6 Conclusion

We have applied three machine learning methods to the classification of TB status utilising demographic and clinical data routinely collected from people who present with a cough at community-level care centres in South Africa. Our experiments were performed using two similar datasets, CODA TB and CAGE-TB. While the former includes a larger number of people, the latter includes a substantially larger set of clinical and demographic data. Using fully nested 10-fold cross validation, we achieved test-set areas under the receiver operating characteristic (AUROCs) of 80.48% and 83.06% for CODA TB and CAGE-TB respectively. We were able to demonstrate improved performance using a one-dimensional convolutional neural network classifier, an architecture not usually used for clinical data. Furthermore, we were able to show that performance improves both when the feature set is extended, and when the dataset increases in size. This holds promise for the development of an easily-applied automated machine-learning based test, that can implemented on a for example a smartphone. Such an implementation could offer improved screening for TB at primary health-care facilities, thereby reducing the incidence of undiagnosed TB and in turn reduce transmission of this disease in the community, especially in developing-world settings.

## Data Availability

All data for CAGE-TB will become available 6-12 months after the project is completed. All data for CODA TB is publicly available online at https://doi.org/10.7303/syn31472953.

## 7 Supplementary

### 7.1 Sequential feature selection

Our experiments include sequential feature selection (SFS), a method that iteratively evaluates feature subsets to identify those most relevant to a particular predictive model. This method is particularly effective in scenarios where the feature space is high-dimensional and domain knowledge about feature importance is limited [Rückstieß et al., 2011]. It operates by incrementally adding features to the feature vector in a greedy fashion, based on their impact on development performance, in our case the AUROC across the nine development sets within the inner-loop. This allows an optimal set of features to be determined. It also provides insight into the relative importance of individual features to the classification process.

Features are always selected employing logistic regression. These optimal features are then applied also to all three classifiers. Because XGBoost ensembles small and highly regularised trees, which already only utilise informative features, it is not itself suitable for feature selection. Similarly, since the kernels impose a minimum input dimensionality for the CNN, it is also not suitable for feature selection. Since the same set of optimised features is used for all three classifiers, performance is directly comparable among the three.

### 7.2 Considered neural network structures

#### 7.2.1 Logistic regression (LR)

Logistic regression is a simple and well-studied binary classifier, that can also be viewed as a single neuron of a multi-layer perceptron with a sigmoid activation function. It computes the posterior probability of the positive class using Equation 1, where *x* is the *d*_feat_-dimensional input feature vector, *w* is the *d*_feat_-dimensional weight vector, *b* is the bias weight, *σ* is the sigmoid activation and *p* is the network output, a score between 0 and 1 that is trained to approximate the posterior probability.

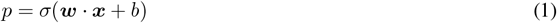

Logistic regression is a linear classifier and therefore has a limited classification ability. It was included primarily as a baseline with which to compare the other, more complex, classifier architectures.

#### 7.2.2 XGBoost

XGBoost is a popular machine learning paradigm which employs gradient boosted ensembles of classification and/or regression trees [Chen and Guestrin, 2016]. This architecture has been shown to be effective when applied to survey-based clinical data [Davagdorj et al., 2020]. Classification trees partition input data based on a set of hierarchical decisions, which are viewed as nodes in a tree structure. Classification is achieved by beginning with the question at the root node and traversing the tree until a leaf node is reached. Tree boosting entails training several weak trees and combining these in an ensemble, referred to as a forest. It is often found that the classification performance of such a forest outperforms a single deep or complex tree. During training, each successive tree is trained to minimise the residual error between the objective and the combined output of the previously-trained trees. As with the other models we considered, binary cross entropy is used as the objective function.

#### 7.2.3 Convolutional neural network (CNN)

We include a convolutional neural network in our evaluation, where this network is configured to receive as input a one-dimensional feature vector, as also used by logistic regression and XGBoost. CNNs are usually configured to input two-dimensional (image) data. To the best of our knowledge, CNNs have not yet been considered for the classification of one-dimensional clinical data, as we do here. Indeed, the concept of convolutional feature detectors, which are well suited to the detection of the localised characteristics in an image, does not seem to be appropriate for the unordered mix of quantitative and categorical data that makes up our dataset. Despite this, we will demonstrate how the CNN is able to achieve the best performance among the three approaches we consider. We hypothesise that this is due to the highly regular structure imposed by the CNN, where only local interactions between inputs are captured (due to the small kernel and its stride) while more global dependencies are captured deeper in the network. When input features that carry related information are in close proximity, the CNN can model this dependency and ignore interactions with features that are further away. We employ the CNN network structure described in Table 8.

**Table 8:** One-dimensional CNN structure. The output dimensions of all but the last layer depend on the input dimension.

| # | Layer type | Kernel dim | Num ($C$) | Stride ($s$) | Act | Layer output |
| --- | --- | --- | --- | --- | --- | --- |
| 1 | Convolution | $(3 \times 1)$ | 20 | 1 | ReLU | - |
| 2 | Max pool | $(2 \times 1)$ | - | 1 | - | - |
| 3 | Convolution | $(3 \times 1)$ | 50 | 1 | ReLU | - |
| 4 | Max pool | $(2 \times 1)$ | - | 1 | - | - |
| 5 | Dense | - | - | - | Sigmoid | 1 |

In preliminary experiments we optimised the following hyperparameters for the CNN architecture using a grid-search within the nested cross validation strategy. Kernel sizes of 2,3 and 5 were tested. Furthermore, we varied the number of CNN channels by considering the combinations [10, 10], [10, 20], [20, 20], [20, 50], [50, 50] and [50, 100], where each value denotes the number of channels for each of the two respective layers. We did not explore deeper models due to small input dimensionality. We also did not explore larger max pooling kernels for the same reason. In fact, we found that for many of the evaluated hyperparameters, model performance was consistent. Taking this into consideration, we elected to utilise the architecture as described in Table 8, which balanced model size, in terms of number of trainable parameters, and performance.

#### Optimisation of CNN feature ordering

Because the kernels in the CNN will capture local dependencies, the ordering of the features in the input vector matters. However, in contrast to an image or a time-domain sequence, the features in the clinical data have no implicit ordering. Therefore we always optimise the ordering by considering 250 randomly-chosen permutations, and select the ordering which achieves the best average development set loss. Finding the optimal ordering from these permutations constitutes 10 · 9 · 250 + 250 = 22750 individual experiments. Due to this computational load, we fix the hyperparameters of the CNN during this search.

#### 7.2.4 Hyperparameter search

Each of the considered classifiers have hyperparameters that must be optimised, as described in Table 9. The table also presents the best hyperparameter values identified during cross-validation, and indicates a large degree of consistency among the two datasets. Note that each of the 10 outer-loop iterations of nested cross validation can result in different hyperparameters.

**Table 9:** Hyperparameters and the ranges considered for optimisation within nested *k*-fold cross validation. Optimal hyperparameters for LR are presented for the subset of features identified during SFS, optimal parameters for the CNN model are presented after further optimisation of the order of the input feature vector identified during SFS. The value for the optimal number of training epochs is calculated as the mean over the outer folds, while modal values are presented for the remainder of the hyperparameters.

| Classifier | Hyperparameter | Range | Optimal CAGE | Optimal CODA |
| --- | --- | --- | --- | --- |
| LR | Learning rate | [0.01, 0.001] | 0.01 | 0.01 |
| LR | Batch size | [8, 16, 32] | 8 | 8 |
| LR | Epochs | [1, ..., 128] | 88 | 59 |
| XGBoost | Max tree depth | [4, 6, 8, 10] | 6 | 6 |
| XGBoost | Min child weight | [1, 4, 8, 10] | 1 | 1 |
| XGBoost | Min split loss | [0, 0.1, 1, 10] | 0 | 0 |
| XGBoost | L2 regularisation | [0, 1, 5] | 1 | 1 |
| XGBoost | L1 regularisation | [0, 1, 5] | 0 | 0 |
| XGBoost | Learning rate | [0.1, 0.3, 0.5, 0.7, 0.9] | 0.3 | 0.3 |
| CNN | Learning rate | [0.01, 0.001] | 0.001 | 0.001 |
| CNN | Batch size | [8, 16, 32] | 32 | 32 |
| CNN | Epochs | [1, ..., 128] | 79 | 120 |

For LR and the CNN, the ADAM optimiser is utilised, with a weight decay of 0.01 Kingma and Ba [2015]. All neural networks were implemented using the open-source machine learning platform PyTorch [Ansel et al., 2024].

### 7.3 Results

#### 7.3.1 CODA Country-wise analysis

Table 10 considers the country-wise performance of the LR model when utilising only the optimal features determined during SFS. It is clear from the table, that there is substantial difference in the model performance on a per country basis, with the worst performance occurring for the Philippines (69.23 ± 3.18%) and the best for India (87.23 ± 1.28%).

**Table 10:**
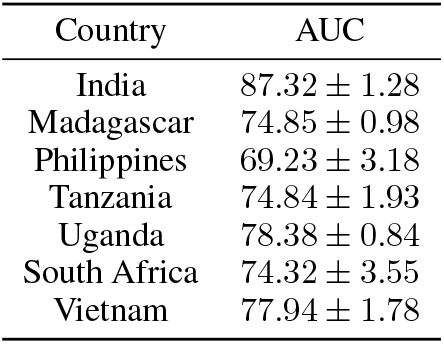
CODA TB country wise development set AUROC (± stddev) for LR.

| Country | AUC |
| --- | --- |
| India | $87.32 \pm 1.28$ |
| Madagascar | $74.85 \pm 0.98$ |
| Philippines | $69.23 \pm 3.18$ |
| Tanzania | $74.84 \pm 1.93$ |
| Uganda | $78.38 \pm 0.84$ |
| South Africa | $74.32 \pm 3.55$ |
| Vietnam | $77.94 \pm 1.78$ |

The effect of each of the individual variables during SFS is presented in Figure 5. It is important to note that the observed absolute improvement or decrease in AUROC may be due to differences in data collection strategies, for example the manner in which the questionnaire data was collected, phrased, and interpreted. It was not feasible in this analysis to systematically evaluate the impact of such factors.

**Figure 5:**
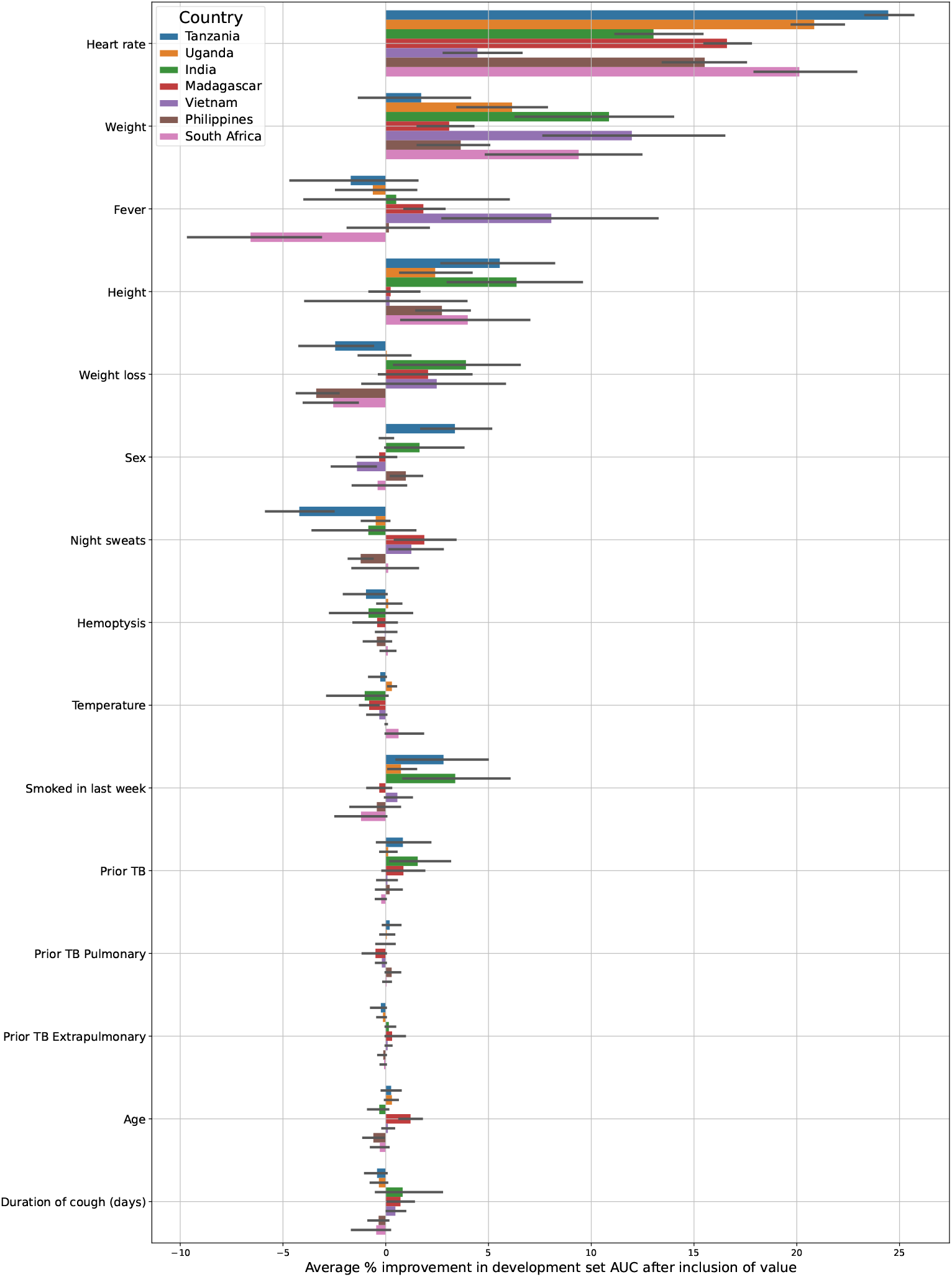
Per-feature average absolute improvement in development AUROC during SFS for the CODA-TB dataset, relative to chance (50% AUROC). Absolute improvement is presented separately for each country, from which AUROC is calculated separately for each country for each of the inner-folds. It is important to note that the average AUROC of the sub-groups is not necessarily equal to the average AUROC of the combined group. Furthermore, for some countries some folds did not contain both positive and negative people. Such cases were omitted from the averages.

For example, from the figure, it appears that including Fever is detrimental to performance for South Africa, while Vietnam benefits greatly from its inclusion. This may suggest that country or site specific classifiers may be necessary to be developed, or indeed that country should be considered as a variable to be input to the classifier.

#### 7.3.2 CNN feature ordering analysis

Because the kernels employed by a CNN capture local patterns, the ordering of the input features affects the effectiveness of the architecture, as described in Section 7.2.3. Figure 6 supports this hypothesis, in which substantial variation in CNN classifier performance is observed when shuffling the order of the features during optimisation.

**Figure 6:**
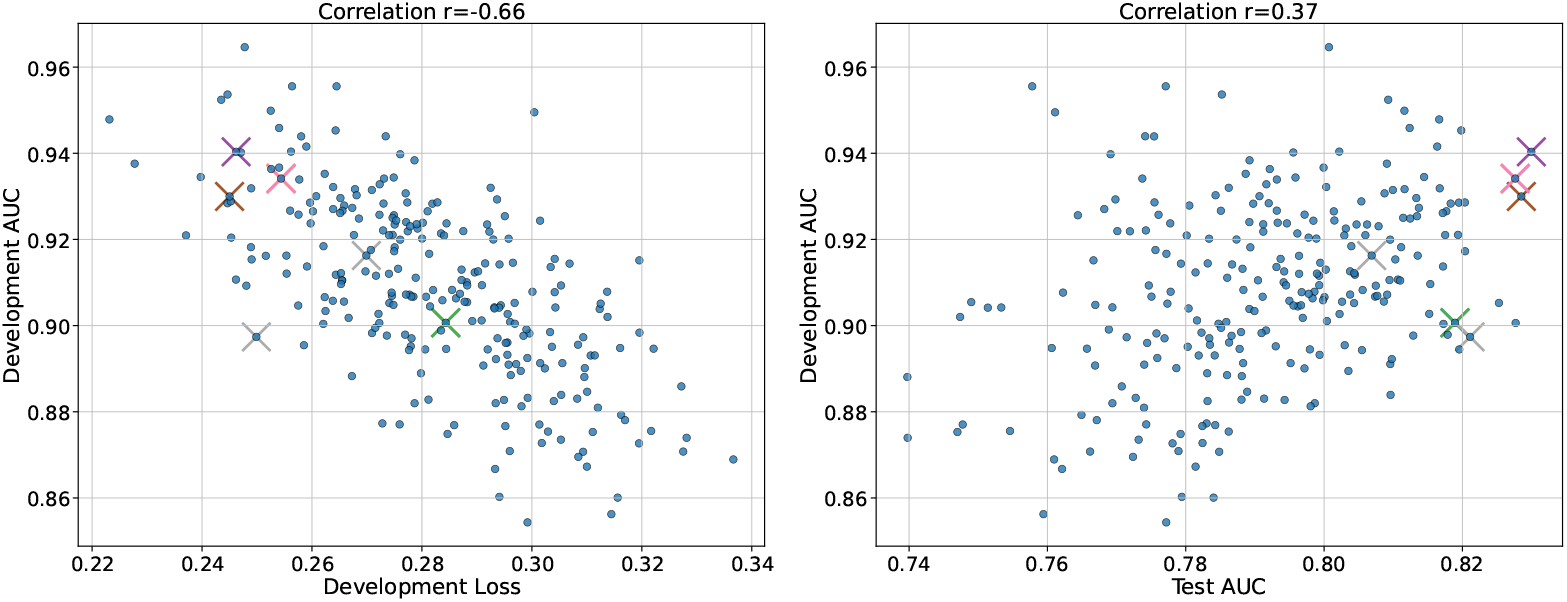
Development loss and test set AUROC versus development set AUROC for each of the 250 considered permutations of the input feature ordering for the CAGE-TB dataset. Pearson correlation coefficients are calculated over these 250 points. Crosses indicate the average of the ordering selected according to the development set results for each of the inner-folds, where each colour indicates a distinct inner-fold. Ideally models should fall into the upper left corner for the left sub-figure and upper right for right sub-figure.

Retraining the CNN using different permutations of the feature order leads to variations in the development set AUROC of up to approximately 10%. We select the optimal ordering by identifying the permutation which minimises the average loss across the development set folds. Although it appears overall that an ordering with a poor development set loss results also in a poor development set AUROC, this relationship is not strong (*r* = − 0.66). Despite this, the optimisation strategy outlined in Section 7.2.3 allows performant vector orderings to be identified and selected, as indicated by the crosses in the figure. Interestingly, for four of the ten folds, exactly the same ordering is identified for the full set of features. When the optimal set of features discovered by SFS is considered, the same ordering is found to be optimal for three of the ten folds.

#### 7.3.3 Ordering results

In Table 11 we present the results after applying the optimisation of the CNN feature vector ordering as described in Supplementary section 7.2.3. For both the CAGE and CODA datasets the shuffling procedure improves both development and test set AUROCs both before and after SFS. Therefore all reported results in the main text are after optimisation of the vector ordering.

**Table 11:**
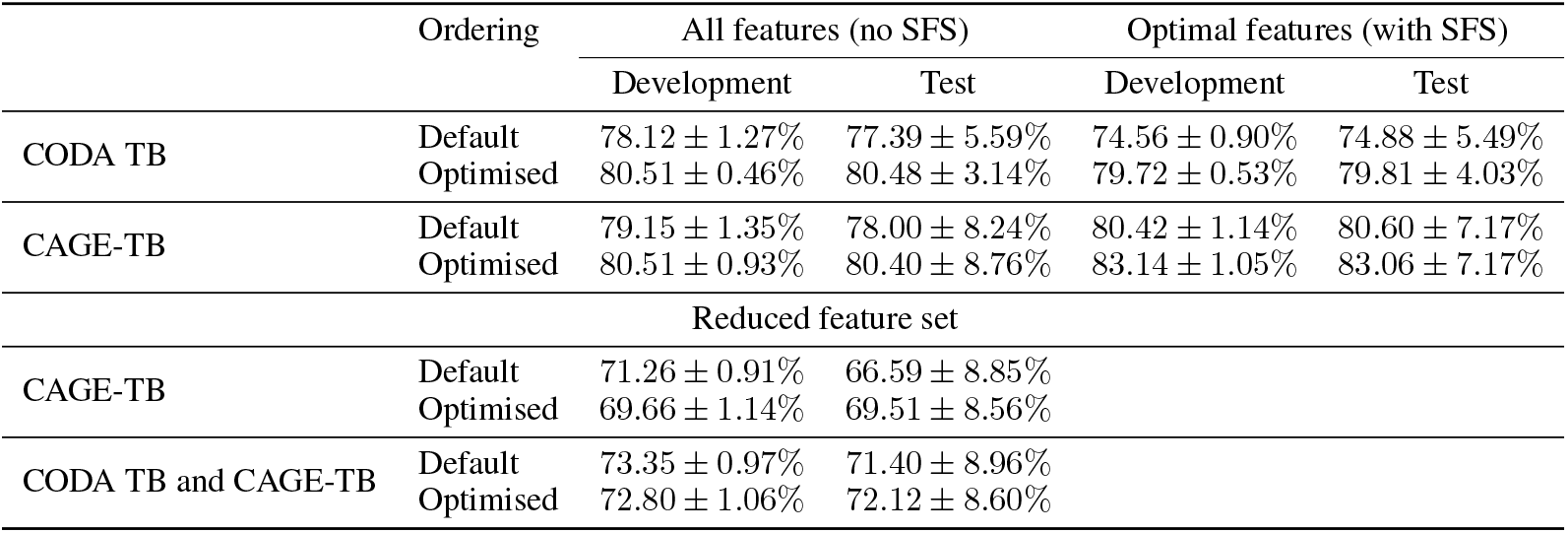
Test-set AUROCs for CNN classifiers trained on either CAGE-TB, CODA TB or merged datasets before and after optimisation of the feature ordering, both when using all 15 features and when using the features identified by SFS.

#### 7.3.4 CNN kernel analysis

Because the input to the classifier is not an image, it is challenging to analyse the learned kernels of the CNN. The left of Figure 7 shows the average test set input vector for TB negative (top) and positive (bottom) people separately, when using the 13 features chosen by SFS in their optimal order. These 13 features constitute a 17 dimensional vector as ART status, CRP and haemoglobin concentration are all multinomial. In the centre of the same figure, the weights of the 20 CNN kernels are shown (*k* = 3 × 1), transposed and arranged as rows, for the first layer. Finally, on the right the average activations for all the negative (top) and positive (bottom) people are presented. One can see that the CNN is learning to consider local interactions in the input layer, as illustrated by the stark contrast both within and between filters. For example, the bottom-most filter outlined in red, is activated when the left two inputs are large. This is true on average for TB positive people who have large values of the inputs *fever* and *heart rate*. This same filter is not activated, on average, for the TB negative people.

**Figure 7:**
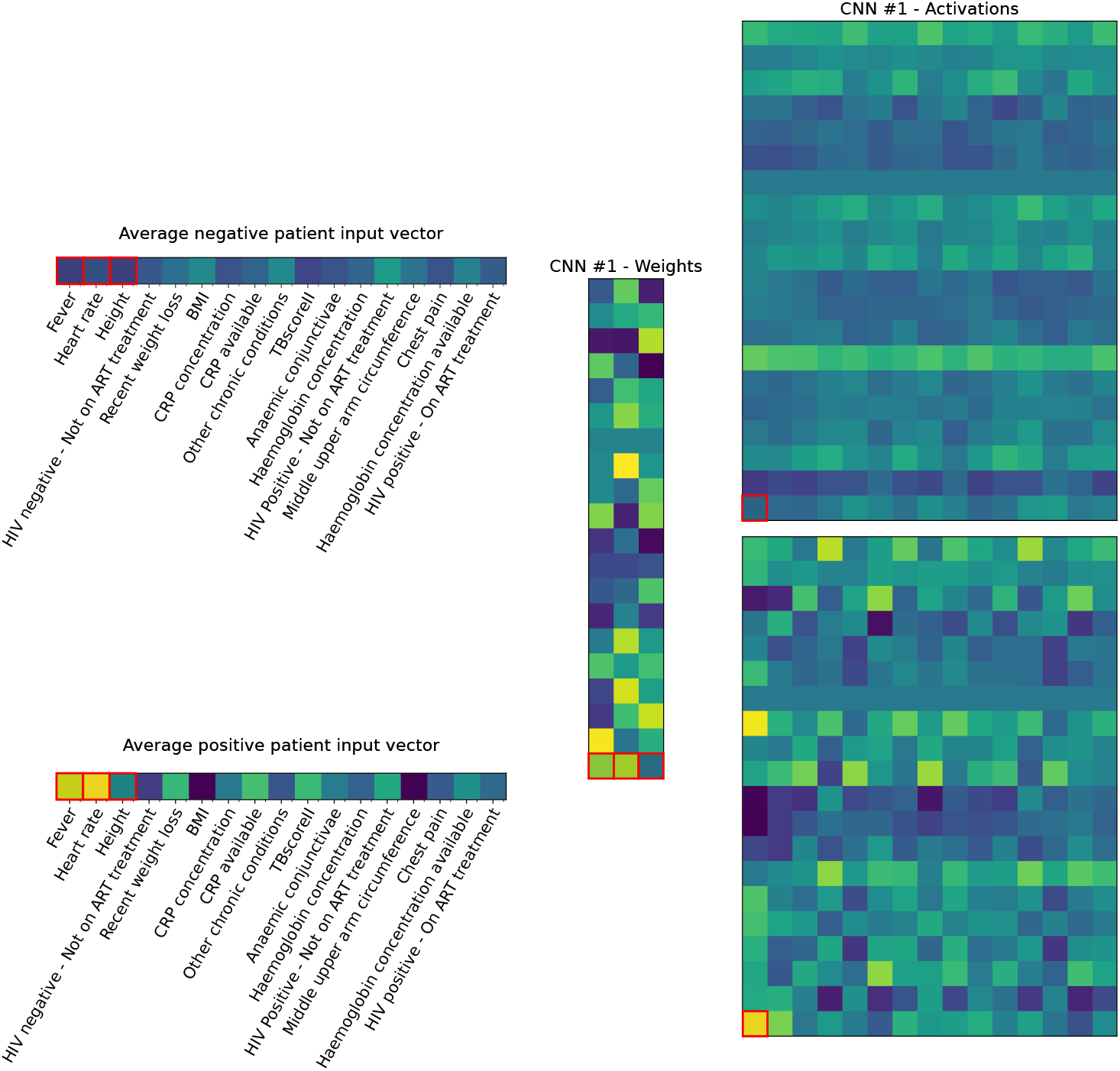
The average test set input vector for TB negative (top left) and positive (bottom left) people, the weights (*k* = 3 × 1) of each of the 20 trained kernels, arranged as rows, and the average activations for the negative (top right) and positive (bottom right) people. Averages are calculated for the first test fold, and the 13 features (17 scalar values) are those chosen from the full set of 38 by SFS, arranged in their optimised order. Lighter (yellow) colours represent large positive values while darker (purple) colours represent negative in terms of magnitude or small values.

**Figure 8:**
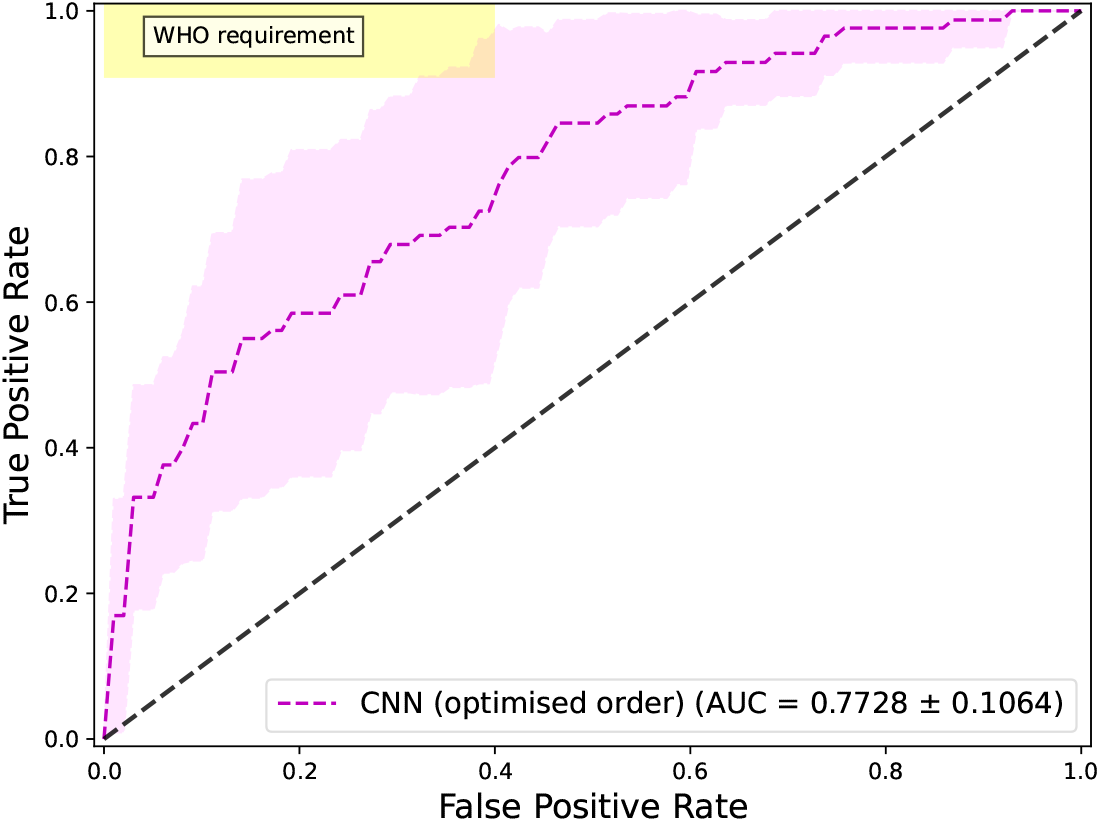
ROC curves reflecting the performance of the convolutional neural network (CNN) classifier when pooling the CODA TB data into the CAGE-TB training data. Results are shown when using the subset of 12 features which occur in both the CAGE-TB and CODA TB datasets.

#### 7.3.5 Merged CNN

### 7.4 Availability experiments results

We additionally conduct SFS for each of the three availability classes separately and in unison, while the SFS for all three classes has been presented in the main text. The three subsequent sub-sections detail noteworthy patters which arise from the feature selection for each of the subsets of availability classes (A, B, A+B). Finally, the development and test set performances for both the CNN and LR classifiers trained on the full subset of features belonging to each class and the input features selected during SFS is presented in Table 12.

**Table 12:** Development and test set AUROC for classifiers trained on the subsets of features separated according to the ease of availability (Table 1) class both on the entire subset and after further optimising the input features using SFS.

| Availability class | Model | All features (no SFS) |  | Optimal features (with SFS) |  |
| --- | --- | --- | --- | --- | --- |
|  |  | Development | Test | Development | Test |
| A | LR | 67.13 ± 1.38% | 67.16 ± 8.01% | 73.31 ± 0.86% | 73.18 ± 7.31% |
| | CNN (Default) | 66.09 ± 1.27% | 66.39 ± 7.57% | 74.06 ± 0.99% | 74.45 ± 8.34% |
| | CNN (Optimised) | 72.82 ± 2.10% | 71.62 ± 11.40% | 75.95 ± 0.83% | 75.88 ± 7.83% |
| B | LR | 79.53 ± 0.98% | 78.93 ± 8.27% | 80.34 ± 0.87% | 80.19 ± 7.62% |
| | CNN (Default) | 77.75 ± 1.21% | 77.30 ± 7.97% | 78.20 ± 1.06% | 78.09 ± 8.32% |
| | CNN (Optimised) | **80.50** ± 1.05% | 79.62 ± 7.35% | 80.53 ± 0.96% | 80.15 ± 6.72% |
| A+B | LR | 77.80 ± 1.23% | 77.85 ± 7.69% | 82.05 ± 0.81% | 81.86 ± 7.08% |
| | CNN (Default) | 78.13 ± 1.16% | 78.02 ± 7.47% | 80.28 ± 1.20% | 79.77 ± 9.05% |
| | CNN (Optimised) | **80.50** ± 0.72% | **79.63** ± 6.73% | **82.13** ± 0.71% | **82.18** ± 5.82% |

#### 7.4.1 Availability class A

In Figure 9 we present the per-feature analysis for LR, as before, but now only consider the subset of features which represent the easiest to collect, which do not rely on measurements which would require a trained health-care practitioner, and are all self-reported. For this subset of features, recent weight loss is the most informative, followed by ART, cough duration, asthma, other chronic conditions, and diabetes. All other values provide no considerable improvement in development set AUROC. As for the feature search applied to the entire set, sex, location, current smoker, arthritis, years since previous TB, dyspnea, previous TB, coughing up sputum, age, fever, night sweats and cholesterol are not found to be useful and degrade classifier performance. Although included in the optimal feature set identified during SFS for CAGE-TB, for the current class of input features, chest pain is no longer informative and leads to a net average reduction in development set AUC.

**Figure 9:**
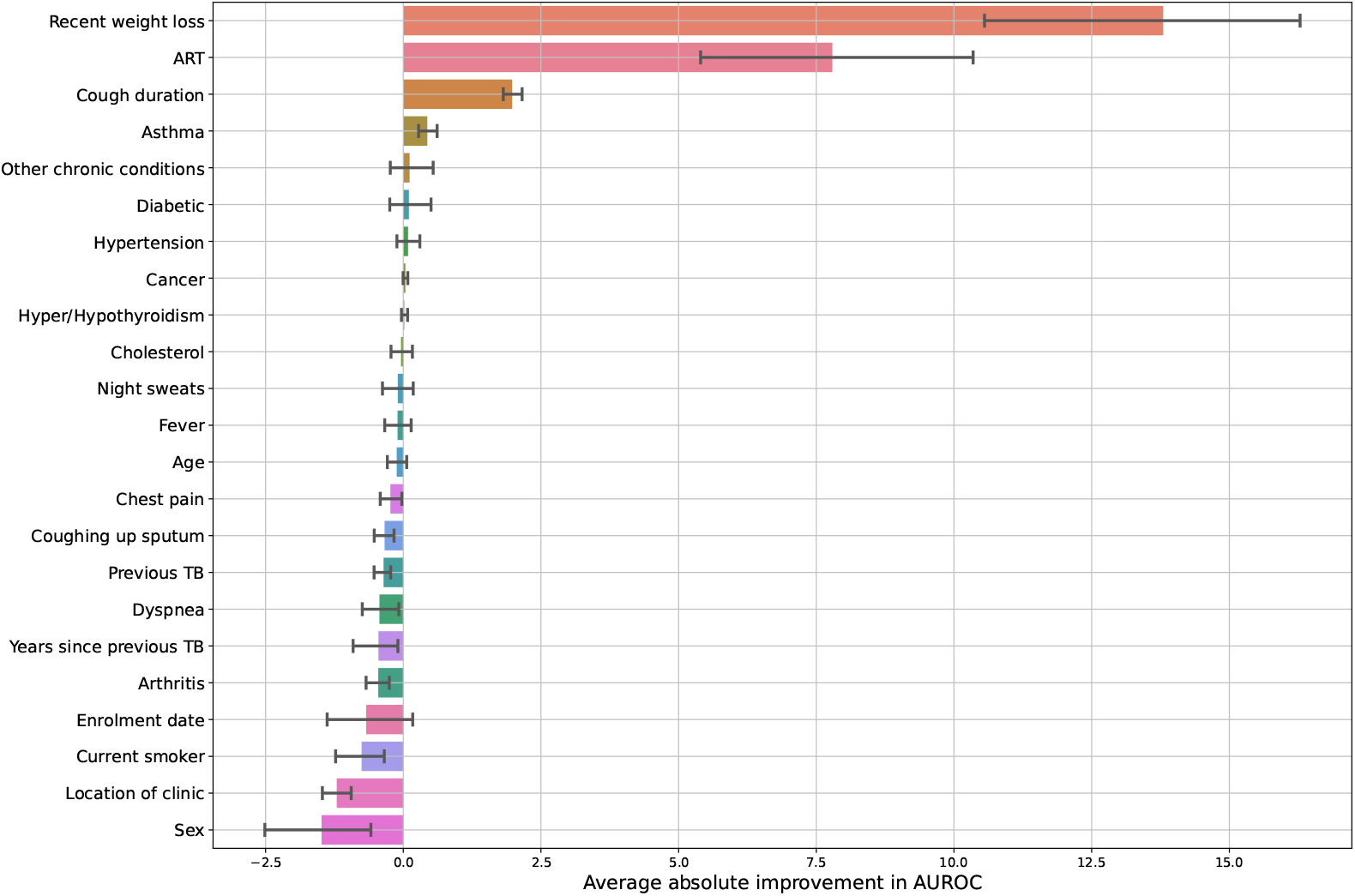
Per-feature average absolute improvement in development set AUROC during SFS for CAGE-TB when only considering the features which belong to the most readily available class A, relative to chance (50% AUROC). Error bars indicate a bootstrap estimate of the 95% confidence interval across the development sets.

#### 7.4.2 Availability class B

Figure 10 presents the feature selection results when applied to the subset of features which are quick to acquire, through some simple measurement but would require the intervention of a health care practitioner are are not purely self-reported, and therefore represent the second class of availability (B). The most informative feature is TBscoreII, which is the second most informative feature overall (only outperformed by CRP concentration). The LR classifier performance improves when additionally including anaemic conjunctivae, middle upper arm circumference, and BMI which all contribute scores to TBscoreII, suggesting that a data centric approach may be able to improve the application of TBscoreII to classification through more nuanced weightings than the integer based scoring strategy. Compared to BMI, height alone found to be more informative whereas weight does not show any performance improvement in SFS. Interestingly, temperature and fagerstrom score are found to be informative, fagerstrom score only marginally, which is not apparent in the feature search which considers all the variables. As for the full feature search, the inclusion of heart rate is found to improve performance. Again, however, HIV, respiratory rate, weight and number of symptoms are not found to improve performance.

**Figure 10:**
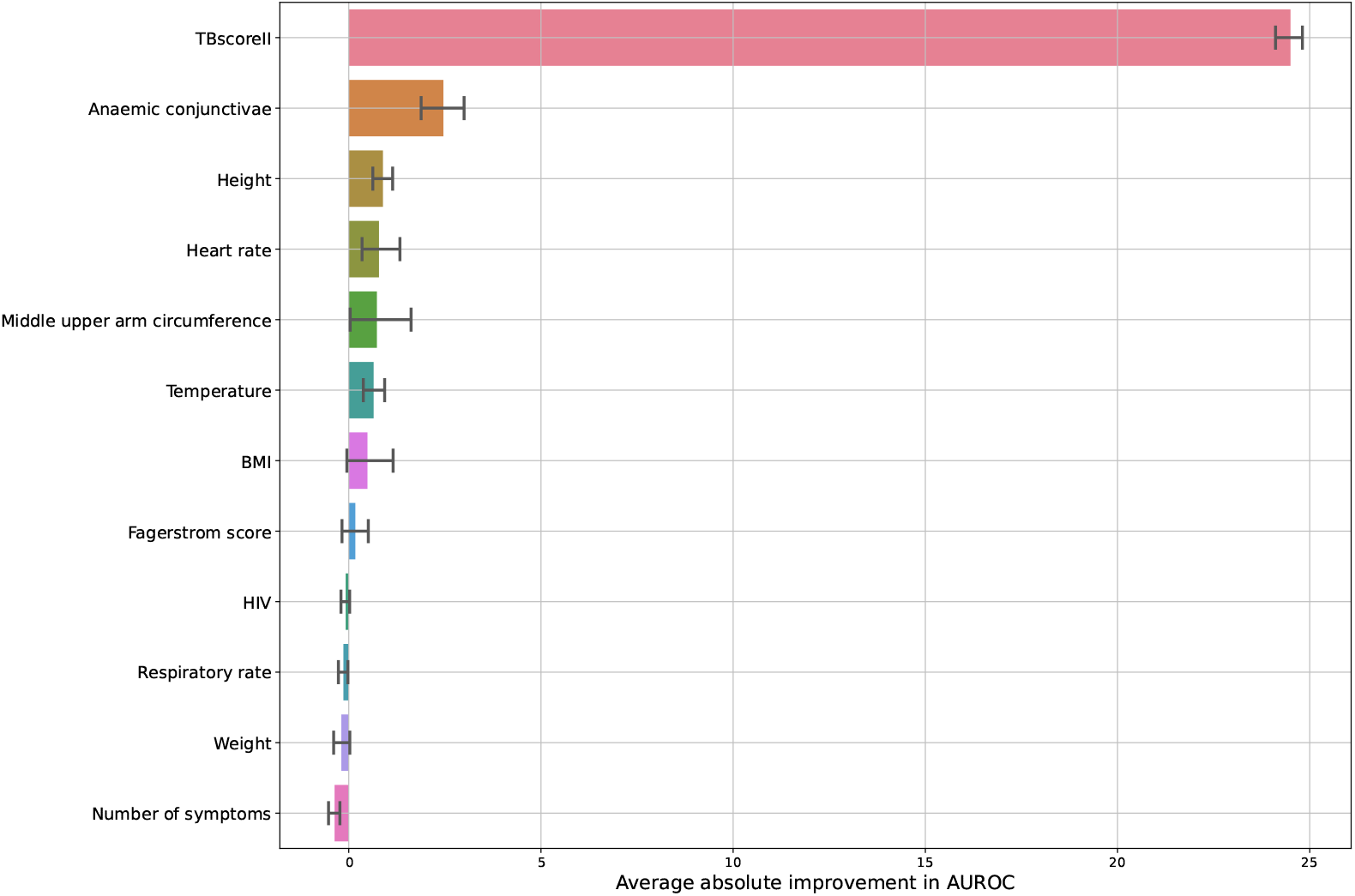
Per-feature average absolute improvement in development set AUROC during SFS for CAGE-TB when only considering the features which belong to availability class B which requires some measurement from a health-care practitioner, relative to chance (50% AUROC). Error bars indicate a bootstrap estimate of the 95% confidence interval across the development sets.

#### 7.4.3 Availability class A and B

Figure 11 presents the feature selection results when considering both the A and B classes of accessibility. As for when considering only class B, TBscoreII is the most informative feature, again followed by anaemic conjunctivae. ART, the next most informative feature, is the first belonging to class A, and as for all previous experiments, is more informative than only knowing whether someone is HIV positive. The subsequent features found to be beneficial to classification performance are fever, height, middle upper arm circumference, BMI, heart rate, temperature.

**Figure 11:**
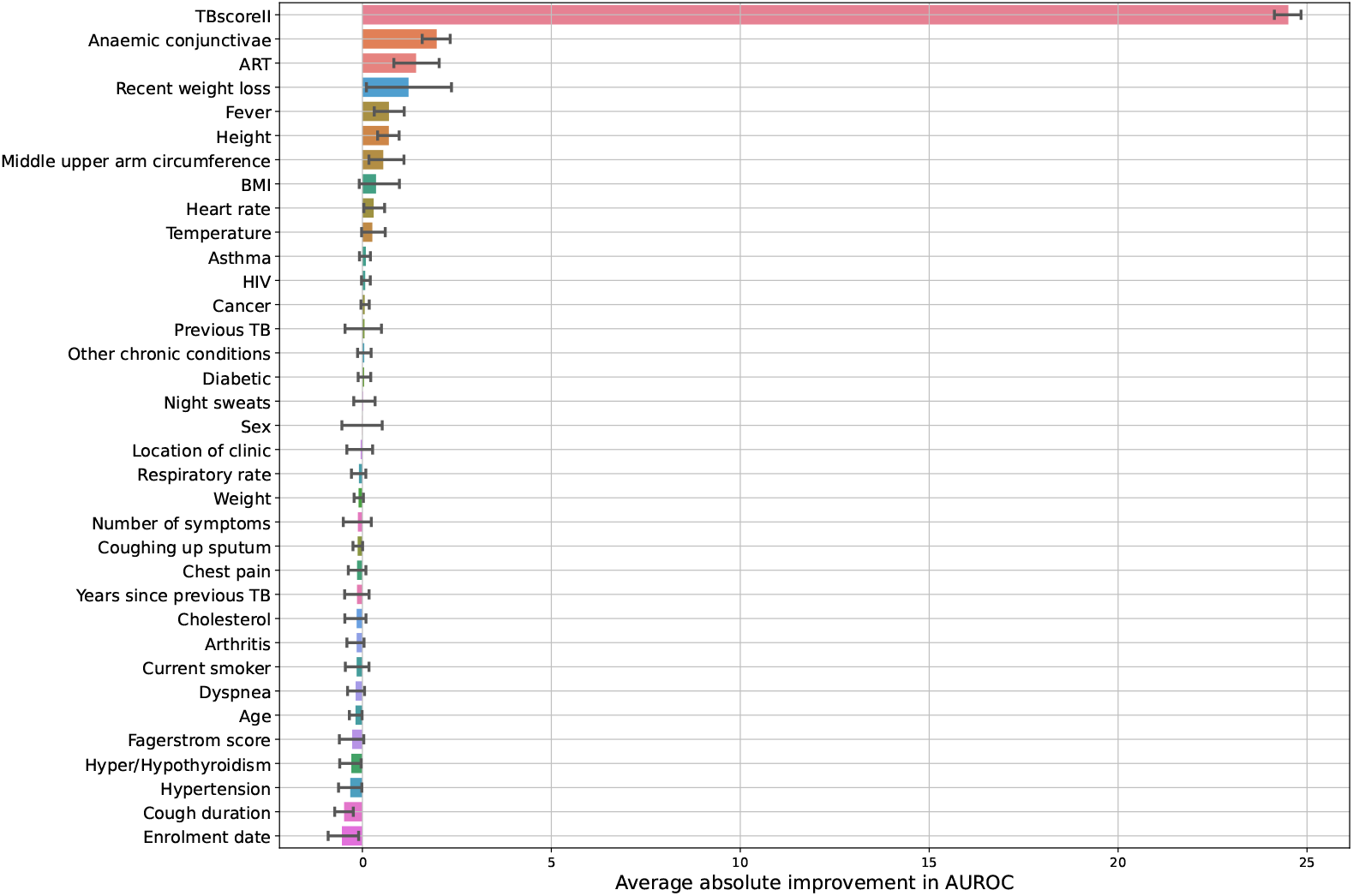
Per-feature average absolute improvement in development set AUROC during SFS for CAGE-TB when only considering the features which belong to both availability class A and B, relative to chance (50% AUROC). Error bars indicate a bootstrap estimate of the 95% confidence interval across the development sets.

Other values such as asthma, HIV, cancer, previous TB, other chronic conditions, diabetes, night sweats provide little to no additional information which improves the ability for the classifier to distinguish TB positives from negatives.

Interestingly, unlike when applying SFS to only the variables in class A, when considering both the values which belong to class A and B, fever becomes 5th most beneficial variable, whereas previously its inclusion lead to an average decrease in AUROC. In contrast, cough duration leads in an average decrease in AUROC whereas previously it become third most beneficial variable.

